# Predictors of Stroke Recurrence After Initial Diagnosis of Cervical Artery Dissection

**DOI:** 10.1101/2024.08.09.24311773

**Authors:** Daniel M. Mandel, Liqi Shu, Christopher Chang, Naomi Jack, Christopher R. Leon Guerrero, Nils Henninger, Jayachandra Muppa, Muhammad Affan, Omair Ul Haq Lodhi, Mirjam R. Heldner, Kateryna Antonenko, David Seiffge, Marcel Arnold, Setareh Salehi Omran, Ross Crandall, Evan Lester, Diego Lopez Mena, Antonio Arauz, Ahmad Nehme, Marion Boulanger, Emmanuel Touze, Joao Andre Sousa, Joao Sargento-Freitas, Vasco Barata, Paulo Castro-Chaves, Maria Teresa Brito, Muhib Khan, Dania Mallick, Aaron Rothstein, Ossama Khazaal, Josefin E. Kaufman, Stefan T. Engelter, Christopher Traenka, Diana Aguiar de Sousa, Mafalda Soares, Sara Rosa, Lily W. Zhou, Preet Gandhi, Thalia S. Field, Steven Mancini, Issa Metanis, Ronen R. Leker, Kelly Pan, Vishnu Dantu, Karl Baumgartner, Tina Burton, Regina von Rennenberg, Christian H. Nolte, Richard Choi, Jason MacDonald, Reza Bavarsad Shahripour, Xiaofan Guo, Malik Ghannam, Mohammad Almajali, Edgar A. Samaniego, Sebastian Sanchez, Bastien Rioux, Faycal Zine-Eddine, Alexandre Poppe, Ana Catarina Fonseca, Maria Fortuna Baptista, Diana Cruz, Michele Romoli, Giovanna De Marco, Marco Longoni, Zafer Keser, Kim Griffin, Lindsey Kuohn, Jennifer Frontera, Jordan Amar, James Giles, Marialuisa Zedde, Rosario Pascarella, Ilaria Grisendi, Hipolito Nzwalo, David S. Liebeskind, Amir Molaie, Annie Cavalier, Wayneho Kam, Brian Mac Grory, Sami Al Kasab, Mohammad Anadani, Kimberly Kicielinski, Ali Eltatawy, Lina Chervak, Roberto Chulluncuy Rivas, Yasmin Aziz, Ekaterina Bakradze, Thanh Lam Tran, Marc Rodrigo Gisbert, Manuel Requena, Faddi Saleh Velez, Jorge Ortiz Gracia, Varsha Muddasani, Adam de Havenon, Venugopalan Y Vishnu, Sridhara Yaddanapudi, Latasha Adams, Abigail Browngoehl, Tamra Ranasinghe, Randy Dunston, Zachary Lynch, Mary Penckofer, James E. Siegler, Silvia Mayer, Joshua Willey, Adeel Zubair, Yee Kuang Cheng, Richa Sharma, João Pedro Marto, Vítor Mendes Ferreira, Piers Klein, Thanh N. Nguyen, Syed Daniyal Asad, Zoha Sarwat, Anvesh Balabhadra, Shivam Patel, Thais Secchi, Sheila Martins, Gabriel Mantovani, Young Dae Kim, Balaji Krishnaiah, Cheran Elangovan, Sivani Lingam, Abid Quereshi, Sebastian Fridman, Alonso Alvarado, Farid Khasiyev, Guillermo Linares, Marina Mannino, Valeria Terruso, Sofia Vassilopoulou, Vasilis Tentolouris, Manuel Martinez Marino, Victor Carrasco Wall, Fransisca Indraswari, Sleiman El Jamal, Shilin Liu, Muhammad Alvi, Farman Ali, Mohammed Sarvath, Rami Z. Morsi, Tareq Kass-Hout, Feina Shi, Jinhua Zhang, Dilraj Sokhi, Jamil Said, Newnex Mongare, Alexis Simpkins, Roberto Gomez, Shayak Sen, Mohammad Ghani, Marwa Elnazeir, Han Xiao, Narendra Kala, Farhan Khan, Christoph Stretz, Nahid Mohammadzadeh, Eric Goldstein, Karen Furie, Shadi Yaghi

## Abstract

**Objective:** Patients with cervical artery dissection (CAD) are at increased ischemic stroke risk. We aimed to identify characteristics that are associated with increased risk of ischemic stroke following initial presentation of CAD and to evaluate the differential impact of anticoagulant versus antiplatelet therapy in these high-risk individuals.

**Methods:** This was a secondary analysis of the Antithrombotic Treatment for Stroke Prevention in Cervical Artery Dissection (STOP-CAD) study, a multicenter retrospective observational study. The primary outcome was subsequent ischemic stroke by day 180 after diagnosis. Patient characteristics were compared between those with vs. without subsequent ischemic stroke. Significant predictors were identified using stepwise Cox regression. Associations between subsequent ischemic stroke risk and antithrombotic therapy type in high-risk patients were explored using adjusted Cox regression.

**Results:** 4,023 patients (mean age 47.4 years; 44.5% were women) were included. By day 180, 5.3% experienced a subsequent ischemic stroke. In adjusted Cox regression, factors associated with increased subsequent ischemic stroke risk were prior ischemic stroke (aHR 7.31, 95% CI 1.61-33.13, p=0.010), presentation within seven days from symptoms, (aHR 3.04, 95% CI 1.04-8.91, p=0.043), infarct on imaging (aHR 9.85, 95% CI 3.65-26.58, p<0.001), and occlusive dissection (aHR 2.34, 95% CI 1.03-5.34, p=0.043). Only patients with occlusive dissection had reduced subsequent ischemic stroke risk with anticoagulation versus antiplatelets (HR 0.37; 95% CI 0.15-0.89, p=0.03).

**Interpretation:** This study identified several predictors of subsequent ischemic stroke among patients with CAD but only patients with occlusive dissection demonstrated a benefit from anticoagulation. These findings require validation by meta-analyses of prior studies.

## Introduction

Cervical artery dissection (CAD) accounts for approximately 2% of all ischemic strokes but emerges as the second-leading cause of stroke in young adults under 45 years of age.^1-3^ The main mechanisms of ischemic stroke after CAD relate to vessel wall injury, which promotes the formation of an intraluminal thrombus or steno-occlusion.^4^ CAD may also cause local symptoms (e.g. neck pain and headache) without ischemic stroke.

The risk of subsequent ischemic stroke is up to 3.5% in randomized clinical trials but as high as 8.7% in observational studies.^5-10^ Moreover, the risk of clinically covert infarcts is substantial, with one study showing that new ischemic brain lesions on MR-imaging occurred in one-quarter of patients in the early phase after diagnosis of CAD.^11^

Antithrombotic treatment (antiplatelets or anticoagulation) is an important aspect of stroke prevention in the management of patients with CAD.^12^ Randomized clinical trials and meta-analyses of observational data have yielded conflicting results regarding the relative effectiveness and safety of anticoagulation compared to antiplatelet treatment.^5-7,13^ The primary results from the recent study, “Antithrombotic Treatment for Stroke Prevention in Cervical Artery Dissection (STOP-CAD)”, showed no significant difference in ischemic stroke risk between anticoagulation versus antiplatelet therapies. Conversely, treatment with anticoagulation up to day 180 significantly increased the risk of major hemorrhage. Nevertheless, exploratory subgroup analyses from the STOP-CAD study found a significant association between anticoagulation use and a lower risk of subsequent ischemic stroke in patients with occlusive dissection.^14^ Identifying patients at higher risk for subsequent ischemic stroke could pinpoint subgroups that may benefit from anticoagulation. In addition, optimizing the triage of these patients to intensified monitoring could potentially influence patient outcomes.

A recent subgroup analysis of the “Aspirin versus anticoagulation in cervical artery dissection (TREAT-CAD)” trial explored whether high-risk clinical features influenced the effects of antithrombotic treatment on the risk of subsequent stroke. The study’s conclusions are constrained by its relatively small sample size and by the use of composite outcomes—both imaging and clinical—instead of exclusively focusing on subsequent ischemic strokes, which limits the specificity of its findings.^15^ Risk factors identified from several observational studies are comparable with those from TREAT-CAD but are similarly limited by sample size.^9,10,16^

Therefore, this study sought to identify clinical features associated with subsequent ischemic stroke (“high risk features”) in CAD patients using a large, contemporary multi-national database, and to evaluate if treatment with anticoagulation was associated with a decreased risk of stroke compared to antiplatelets.

## Methods

### Institutional review board approval

This analysis was conducted in accordance with the ethical guidelines of the original STOP-CAD study, which received Institutional Review Board (IRB) approval. As this is a secondary analysis of de-identified data, additional IRB approval was waived by the Lifespan IRB. Aggregate data will be shared upon reasonable request to the STOP-CAD principal investigator.

### Study design and patient population

We conducted a pre-planned secondary analysis of the STOP-CAD study. The STOP-CAD study was a multicenter international retrospective observational study of patients with non-major trauma-related CAD. Methodological details from STOP-CAD have been previously described.^14^ The STOP-CAD study aimed to compare the safety and efficacy of antiplatelets to anticoagulation. All patients from the STOP-CAD study were included in this analysis.

### Study variables

Selected variables reported for the STOP-CAD study were used; further details are provided in the main paper.^14^ For the present study, we collected the following co-variates:

#### Demographics

age, biological sex, race (White, Black, Asian, Other/Unknown), and ethnicity (Hispanic vs. non-Hispanic).

#### Clinical variables

history of migraine, history of hypertension, history of diabetes, history of hyperlipidemia, active smoking, known history of connective tissue disease, a history of prior ischemic stroke, time from first symptoms to dissection diagnosis (0-7 days vs. > 7 days), and recent history of non-major neck trauma.

#### Imaging variables

infarct on baseline imaging (CT or MRI), intracranial dissection or extension, location of dissection (carotid, vertebral, multivessel, other/undetermined), partially occlusive intraluminal thrombus at or distal to the dissection site, intracranial large vessel occlusion, occlusive dissection (defined as complete occlusion at the dissected artery), and pseudoaneurysm.

#### Treatment arms

antiplatelet (single or dual) and anticoagulation (therapeutic parenteral anticoagulation with heparin or low molecular weight heparin or oral anticoagulation with direct oral anticoagulation or vitamin K antagonist).

### Study Outcome

The study outcome was subsequent ischemic stroke by day 180 after initial diagnosis. Subsequent ischemic stroke was defined as new or worsening neurologic symptoms referable to the territory of the dissected artery and either (1) lasting for at least 24 hours or (2) with imaging evidence of new or enlarging acute infarction.

### Analytical Plan

The patient cohort was divided into two groups based on the occurrence of a subsequent ischemic stroke by day 180 after diagnosis. Patient demographic, clinical, and imaging variables were compared between groups to identify potential predictors of subsequent stroke. Chi-square test, Fisher exact test, Wilcoxon Rank Sum, and Student t-test were conducted as appropriate. Univariable analyses were performed to preliminarily assess each variable’s association with the incidence of subsequent ischemic stroke, applying a p-value cutoff of <0.2 to select variables for further multivariable stepwise Cox regression. Lasso regression was executed to confirm the most important predictors. In further analysis, adjusted Cox regression models were used to explore the impact of anticoagulation versus antiplatelet therapy on patients with high-risk predictors that reached statistical significance in the preceding multivariable stepwise Cox regression. Kaplan-Meier curves were constructed to depict event rates across treatment arms for each high-risk subgroup. In these models, patients were categorized based on the initial treatment they received—either anticoagulation or antiplatelet—and the follow-up period started on the day of dissection diagnosis. Censoring was applied at the occurrence of a subsequent ischemic stroke, at the last known follow-up until day 180, upon death, upon switching between anticoagulation and antiplatelet treatments or cessation of therapy, or after stent placement at dissection site. Outcomes counted towards the treatment the patient was on at the time of occurrence. Data were analyzed using StataSE 18.

## Results

The analysis included 4,023 patients with an average age of 47.4 years and 44.5% of the population were women. The rate of subsequent ischemic stroke was 5.3% (214/4023) by 180 days. The rate of subsequent ischemic stroke that occurred by day 7, day 15, and day 30 was 3.0% (122/4023), 4.0% (160/4023), and 4.6% (187/4023), respectively.

### Factors associated with subsequent ischemic stroke risk in univariable analyses

In univariable analyses, factors associated with an increased risk of subsequent ischemic stroke were older age (mean age: 49 vs. 46 years old, p=0.042), hyperlipidemia (29.0% vs. 22.6%, p=0.030), prior history of ischemic stroke (7.5% vs. 4.2%, p=0.024), presentation within seven days from symptom onset (83.6% vs. 75.8%, p=0.009), acute infarct on baseline imaging (77.1% vs. 57.0%, p<0.001), occlusive dissection (47.2% vs. 35.8%, p=0.001), and intracranial large vessel occlusion (34.1% vs. 23.6%, p<0.001; Table 1).

**Table 1.** Univariable Analysis of Preselected Variables Across Patients With or Without Subsequent Ischemic Stroke.

|  | **Subsequent ischemic stroke**<br>**(n = 214)** | **No subsequent ischemic stroke**<br>**(n = 3809)** | **p-value** |
| --- | --- | --- | --- |
| <u>Demographics</u> |  |  |  |
| **Age (median, IQR)** | 49 (39-58) | 46 (38-56) | 0.042 |
| **Sex (% Women)** | 83 / 214 (38.8%) | 1709 / 3809 (44.9%) | 0.082 |
| **Race** |  |  | 0.809 |
| **White** | 152 / 214 (71.0%) | 2800 / 3809 (73.5%) |  |
| **Black** | 15 / 214 (7.0%) | 227 / 3809 (6.0%) |  |
| **Asian** | 8 / 214 (3.7%) | 141 / 3809 (3.7%) |  |
| **Other or unknown** | 39 / 214 (18.2%) | 641 / 3809 (16.8%) |  |
| **Hispanic Ethnicity** | 177 / 190 (93.2%) | 3132 / 3455 (90.7%) | 0.245 |
| <u>Clinical History</u> |  |  |  |
| **Migraine** | 37 / 214 (17.3%) | 631 / 3809 (16.6%) | 0.782 |
| **Hypertension** | 80 / 214 (37.4%) | 1344 / 3809 (35.3%) | 0.532 |
| **Diabetes** | 18 / 214 (8.4%) | 322 / 3809 (8.5%) | 0.983 |
| **Hyperlipidemia** | 62 / 214 (29.0%) | 859 / 3809 (22.6%) | 0.030 |
| **Active smoking** | 47 / 214 (22.0%) | 736 / 3809 (19.3%) | 0.343 |
| **Connective tissue disease** | 3 / 214 (1.4%) | 80 / 3809 (2.1%) | 0.626 |
| **History of prior ischemic stroke** | 16 / 214 (7.5%) | 161 / 3809 (4.2%) | 0.024 |
| **Presentation within 7 days of first symptoms** | 179 / 214 (83.6%) | 2871 / 3787 (75.8%) | 0.009 |
| **Non-major neck trauma** | 57 / 214 (26.6%) | 835 / 3808 (21.9%) | 0.107 |
| <u>Imaging</u> |  |  |  |
| **Acute infarct on initial imaging** | 165 / 214 (77.1%) | 2171 / 3809 (57.0%) | <0.001 |
| **Intracranial dissection or extension** | 23 / 214 (10.7%) | 511 / 3809 (13.4%) | 0.263 |
| **Dissection location** |  |  | 0.207 |
| **Carotid dissection only** | 107 / 214 (50.0%) | 1903 / 3809 (50.0%) |  |
| **Vertebral dissection only** | 76 / 214 (35.5%) | 1479 / 3809 (38.8%) |  |
| **Other intracranial vessel or unknown location** | 2 / 214 (0.9%) | 13 / 3809 (0.3%) |  |
| **Multivessel dissection** | 29 / 214 (13.6%) | 414 / 3809 (10.9%) |  |
| **Partially occlusive thrombus** | 28 / 214 (13.1%) | 381 / 3809 (10.0%) | 0.147 |
| **Intracranial large vessel occlusion** | 73 / 214 (34.1%) | 900 / 3809 (23.6%) | <0.001 |
| **Occlusive dissection** | 100 / 212 (47.2%) | 1351 / 3769 (35.8%) | 0.001 |
| **Pseudoaneurysm** | 20 / 214 (9.3%) | 527 / 3809 (13.8%) | 0.062 |

IQR = interquartile range

### Predictors of subsequent ischemic stroke in stepwise multivariable Cox regression

In the stepwise multivariable Cox regression analysis, which included variables with a p-value <0.2 from univariate analyses, factors associated with a higher risk of subsequent ischemic stroke were prior history of ischemic stroke (aHR 7.31, 95% CI 1.61-33.13, p=0.010), presentation within 7 days from symptom onset (aHR 3.04, 95% CI 1.04-8.91, p=0.043), acute infarct on baseline imaging (aHR 9.85, 95% CI 3.65-26.58, p<0.001), and occlusive dissection (aHR 2.34, 95% CI 1.03-5.34, p=0.043; Table 2). The Lasso regression suggested that the presence of infarct on imaging was the most contributing predictor in the multivariable model.

**Table 2.** Unadjusted and Adjusted Cox Regression Comparing Hazard Ratios for Statistically Significant Variables Identified in Multivariable Stepwise Cox Regression.

|  | **Unadjusted HR 95% CI p-value** | **Adjusted HR 95% CI p-value** |
| --- | --- | --- |
| **Prior history of ischemic stroke** | HR 7.60, 95% CI 1.66 - 34.84, p=0.009 (*) | aHR 7.31, 95% CI 1.61 - 33.13, p=0.010 (*) |
| **Presentation within 7 days from first symptoms** | HR 1.72, 95% CI 1.19 - 2.48, p=0.004 | aHR 3.04, 95% CI 1.04 - 8.91, p=0.043 (*) |
| **Infarct of baseline imaging** | HR 2.55, 95% CI 1.84 - 3.54, p<0.001 | aHR 9.85, 95% CI 3.65 - 26.58, p<0.001 (*) |
| **Occlusive dissection** | HR 4.46, 95% CI 1.98 - 10.01, p<0.001 (*) | aHR 2.34, 95% CI 1.03 - 5.34, p=0.043 (*) |

HR = Hazard Ratio, aHR = adjusted Hazard Ratio, CI = confidence Interval.
(\*) Failed Schoenfeld proportional hazard assumption test; accelerated failure time model used instead.

### The benefit of anticoagulation based on factors associated with ischemic stroke

Among patients with the high-risk features above, those with occlusive dissection had a significantly lower risk of subsequent ischemic stroke with anticoagulation compared to antiplatelet treatment (HR 0.37, 95% CI 0.15-0.89, p=0.03; Figure 1a). This was not the case with the other predictors of subsequent ischemic stroke: prior history of ischemic stroke (HR 1.78, 95% CI 0.46-6.85, p=0.40; Figure 1b), presentation within seven days of symptom onset (HR 0.79, 95% CI 0.38-1.64, p=0.53; Figure 1c), and acute infarct on baseline brain imaging (HR 0.85, 95% CI 0.41-1.78, p=0.67; Figure 1d).

**Figures 1a-d.**
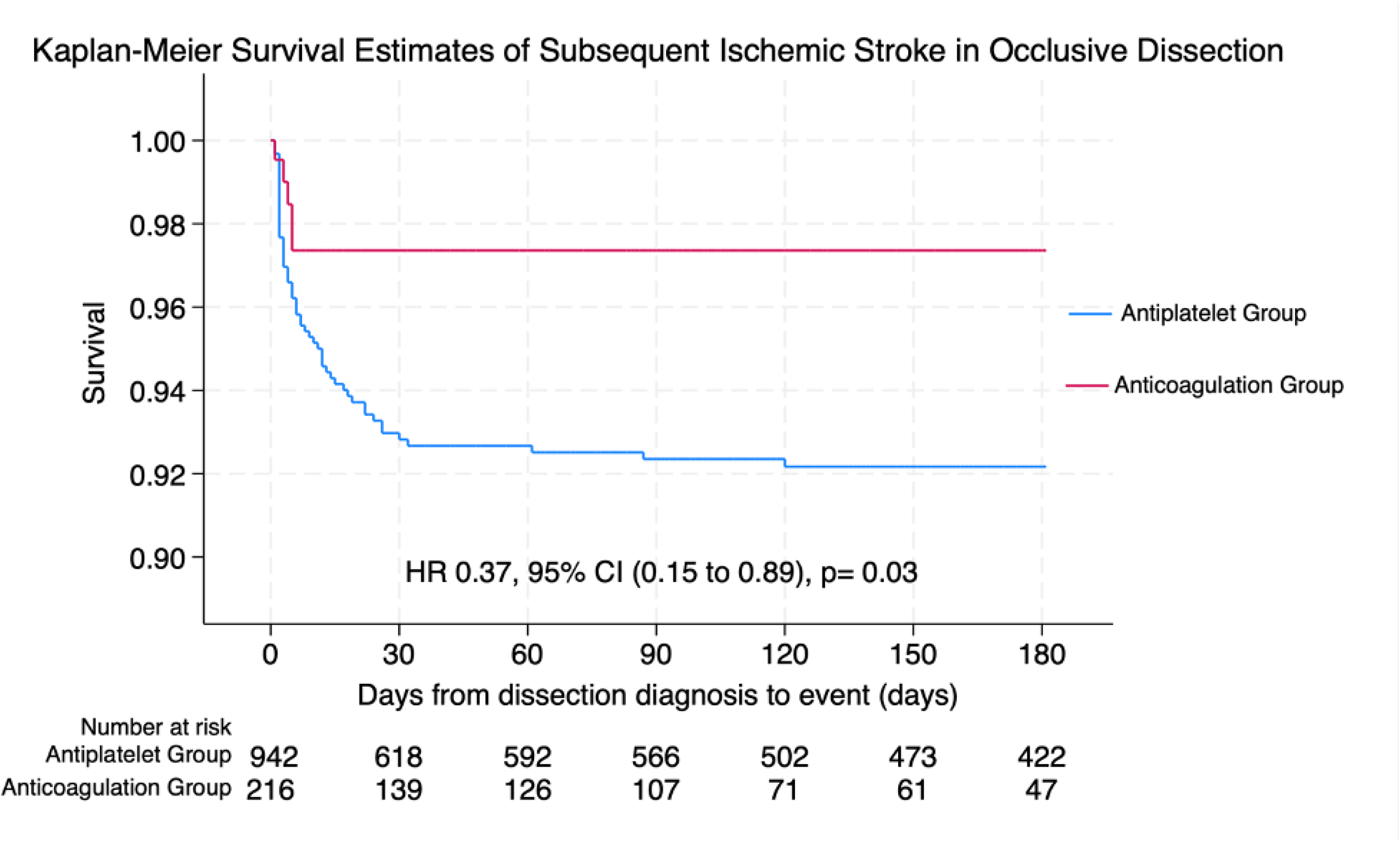
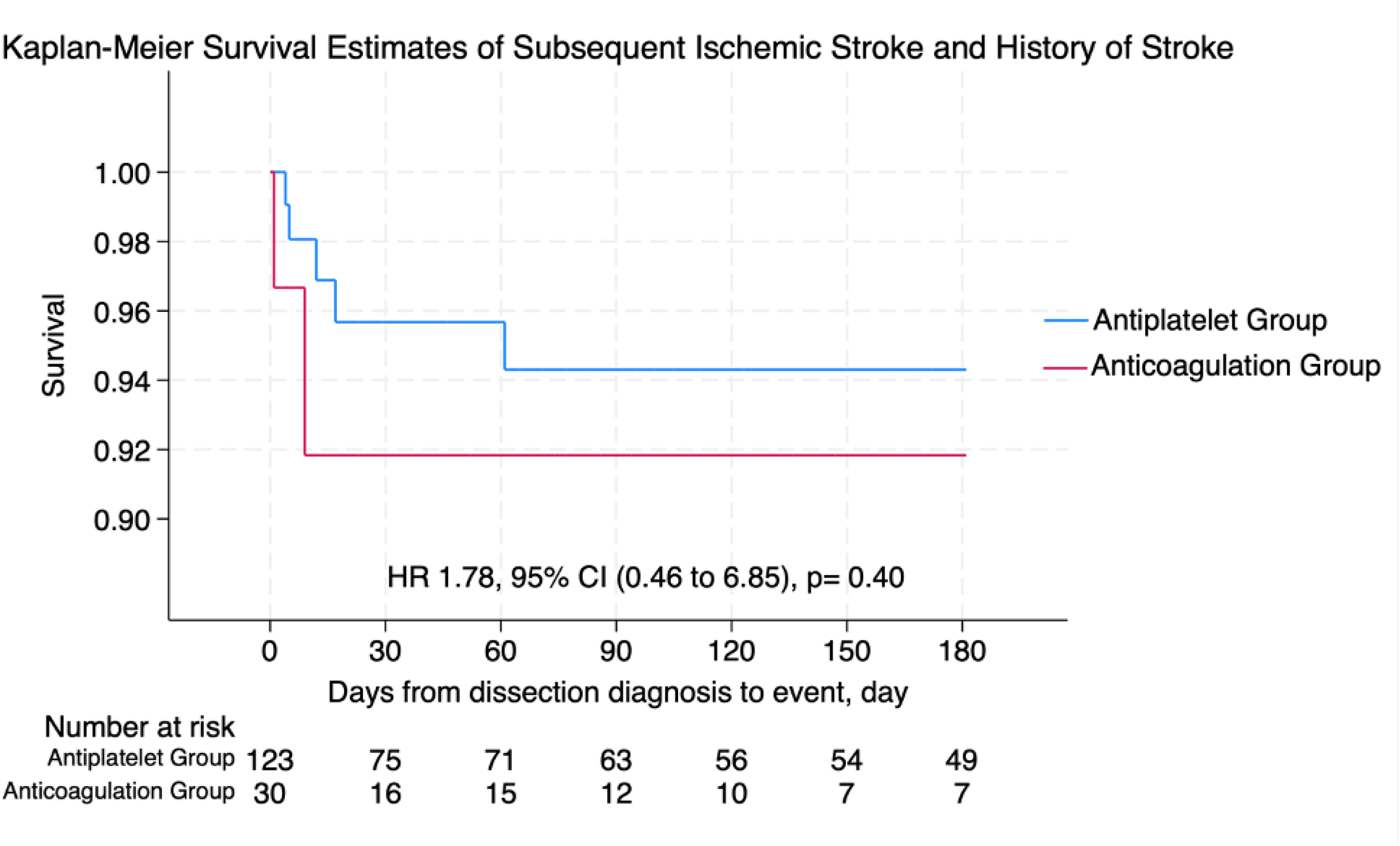
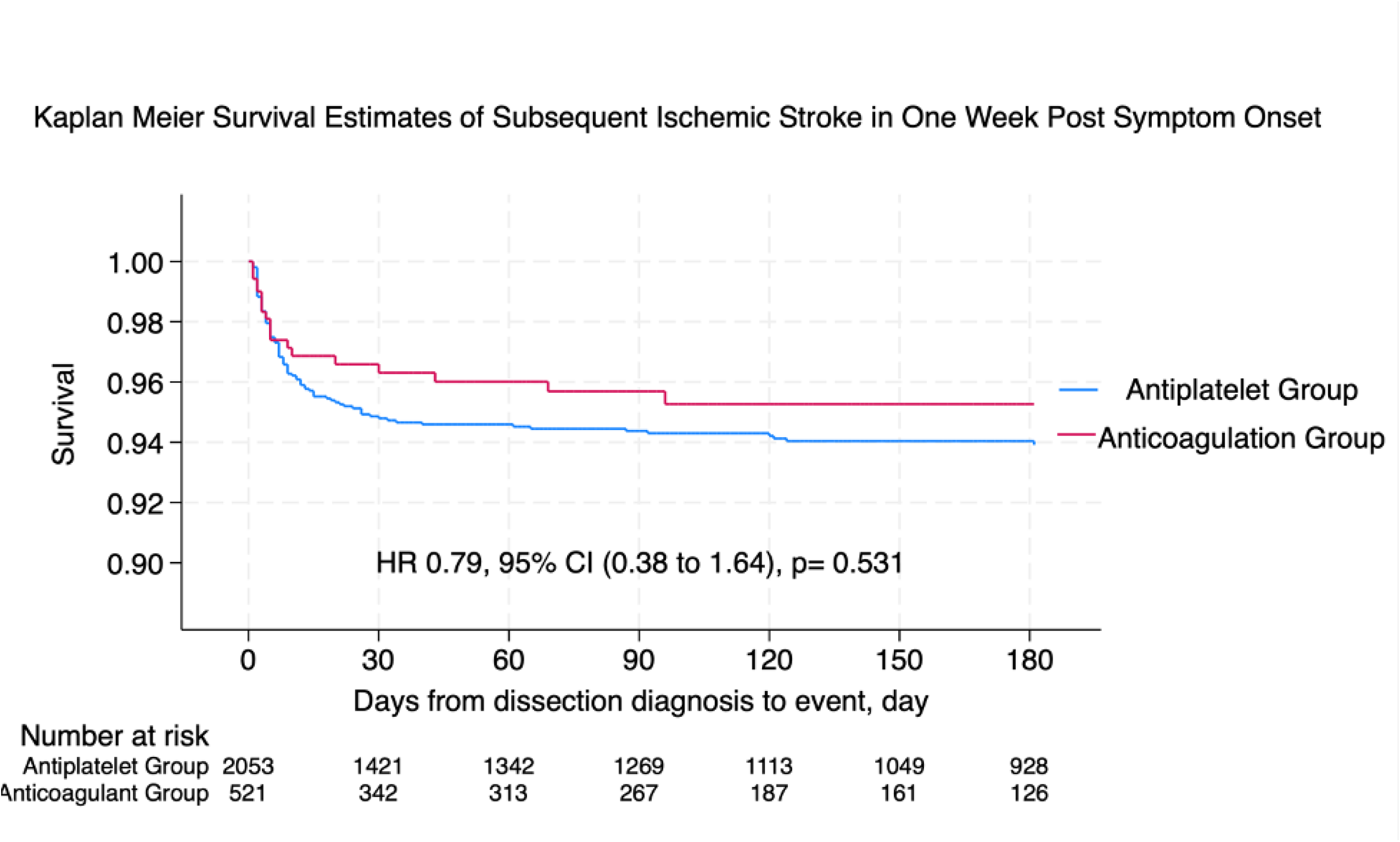
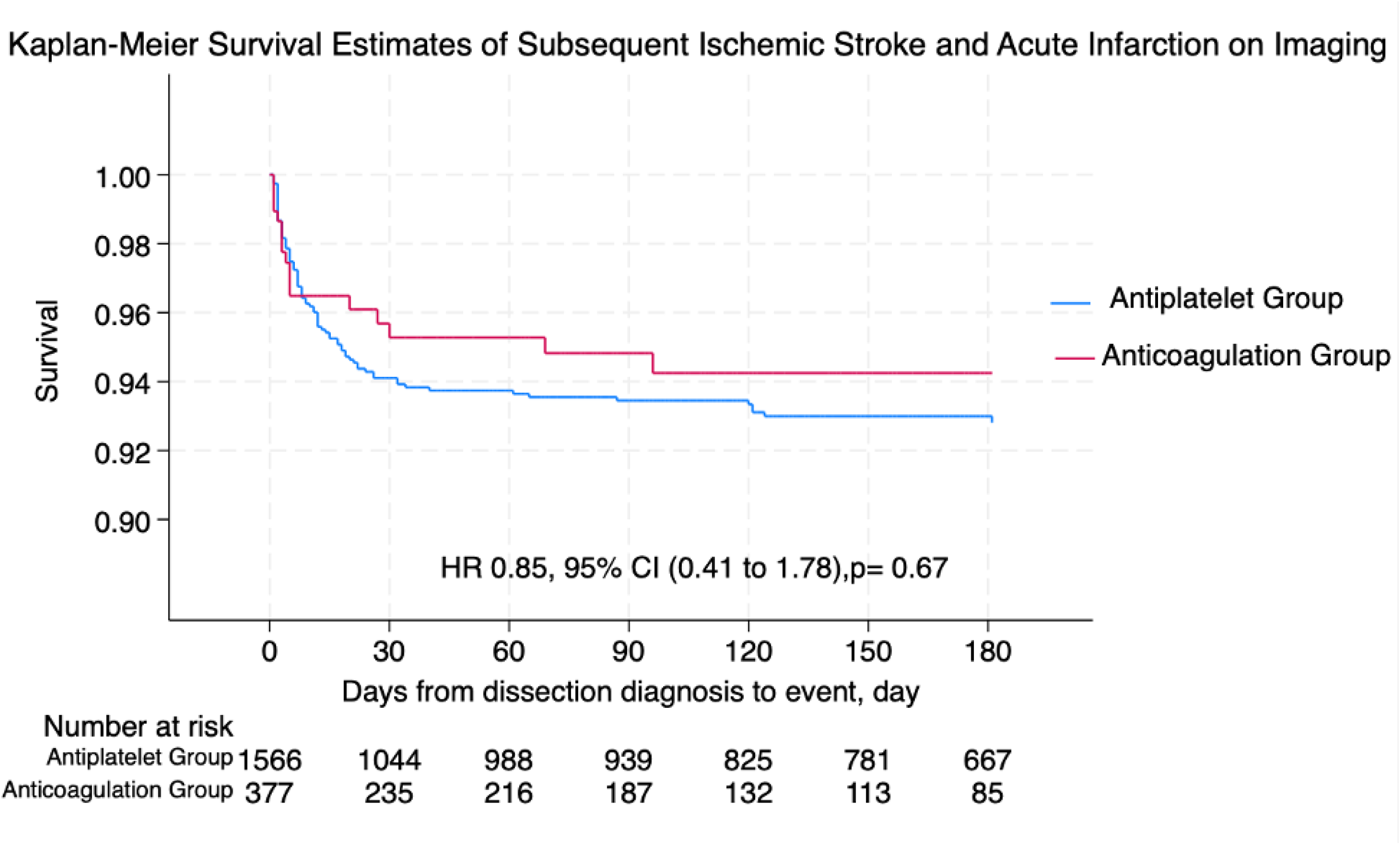
Kaplan-Meier Estimates comparing subsequent ischemic stroke event rates with anticoagulation versus antiplatelets in high-risk subgroups.

## Discussion

This post-hoc analysis of the STOP-CAD study found that among patients diagnosed with acute non-major trauma-related CAD, high-risk features for a subsequent stroke following the index presentation included a prior history of ischemic stroke, presentation within seven days of dissection symptom onset, acute infarct on initial imaging, and occlusive dissection. Among these high-risk features, acute infarct on initial imaging was identified as the most important factor associated with subsequent ischemic stroke, in keeping with prior studies.^4,7^

Several studies have investigated predictors of subsequent ischemic stroke after initial diagnosis of CAD.^5-10,15^ The TREAT-CAD trial found that the risk of stroke was highest within two weeks of randomization.^7^ Other observational studies showed that multivessel dissection, early recurrent dissection, occlusive dissection, and cerebrovascular ischemia on presentation were associated with increased risk of ischemic stroke.^8-10,16^ Recently, the American Heart Association published a scientific statement regarding the treatment for CAD.^12^ In this article, the predictors of ischemic stroke present in greater than one reviewed study included male sex, current smoking, vertebral artery involvement, multiple dissections, early recurrent dissection, high-grade stenosis or occlusion, and intraluminal thrombus.^12^ Consistent with several of the aforementioned observations, the current study found that early presentation, cerebrovascular ischemia, and occlusive dissection were associated with increased risk of subsequent ischemic stroke. In contrast, male sex, smoking, vertebral artery involvement, multivessel dissections, and presence of an intraluminal thrombus were not significantly associated with an increased risk of subsequent ischemic stroke.

Occlusive dissection is a variable of particular interest in this study given the implications for a particular therapeutic strategy. This study found that patients with occlusive dissection who were treated with anticoagulation had a decreased risk of subsequent ischemic stroke compared to those treated with antiplatelets. This was not the case for other high-risk predictive factors identified in this study.

Prior studies suggested a benefit of anticoagulation over antiplatelet in patients with carotid occlusion,^17^ possibly related to reducing further thrombosis and stump embolization which frequently occurs in the early phase following index presentation.^18^ Likewise, the current study illustrates the benefit of anticoagulation in patients with occlusive CAD, which may lower the risk of subsequent ischemic stroke by a similar mechanism. Other predictors of subsequent ischemic stroke in this study do not seem to have a plausible mechanistic benefit from anticoagulation.

In the present study, there was no association between intraluminal thrombus and subsequent ischemic stroke risk. In the STOP-CAD study, patients with intraluminal thrombus were more likely to be treated with anticoagulation,^14^ potentially reducing their risk. Additionally, the limited number of patients with intraluminal thrombus precluded definitive conclusions about the relative risk this predictor poses or the efficacy of anticoagulation compared to antiplatelet therapy in this specific group. Although this study showed a marginally increased risk of subsequent ischemic stroke in patients with intraluminal thrombus (13.1%) versus those without (10.0%), the difference was not statistically significant (p = 0.147; Table 1), possibly reflecting the small sample size of patients with this feature. Thus, larger studies or meta-analyses of existing studies are needed to further explore this finding.

Notably, results from the recently published subgroup analysis of the TREAT-CAD trial suggested that anticoagulation might be a more effective treatment in CAD patients with *non-occlusive* dissection. This conclusion was supported by the finding that patients without occlusion had decreased odds of the study’s combined clinical and radiographic outcomes when treated with anticoagulation compared to those treated with aspirin.^15^ The noticeable divergence between the current study’s findings and those from the TREAT-CAD study could arise from the current study’s exclusive focus on ischemic stroke as the primary endpoint, in contrast to the TREAT-CAD study, which evaluated a composite of clinical and radiographic outcomes, encompassing both ischemic and hemorrhagic events. Accordingly, when isolating clinically-apparent ischemic stroke outcomes in the TREAT-CAD subgroup analysis, subsequent ischemic strokes were numerically higher in the antiplatelet group among those with occlusive dissection (3/32; 9.4%) compared to those with non-occlusive dissection (4/58; 6.8%).

No ischemic strokes occurred in the vitamin K antagonist group among those with occlusive dissection (0/23) and non-occlusive dissection (0/59).^15^ Despite the limited sample size in the TREAT-CAD subgroup analysis, the clinical outcome data suggests that the beneficial effect of anticoagulation is at least numerically more pronounced in those with occlusive dissection versus non-occlusive dissection. It is within this framework that the seemingly disparate findings from the two studies can be reconciled.

This study has several limitations. First, the retrospective and observational nature of the STOP-CAD study may affect the findings through confounding by indication, particularly when comparing antiplatelet to anticoagulation across high-risk sub-groups, although it is conceivable that the assumed influence would bias against anticoagulation and likely result in a reduced treatment effect of anticoagulation in high-risk sub-groups. This is particularly the case in patients with partially occlusive intraluminal thrombus where anticoagulation is generally preferred. Second, the patient cohort, primarily drawn from tertiary care centers at major academic institutions with high rates of mechanical thrombectomy, might not represent the full spectrum of stroke severity. Therefore, the findings from the present study may not fully extend to individuals with milder strokes or localized symptoms or to patients managed in smaller, non-academic healthcare settings. Third, the inclusion of the variable ‘prior history of ischemic stroke’ in this analysis has interpretive limitations, as the specific causes and timing of previous strokes were not determined. Fourth, the comparisons of anticoagulants and antiplatelets within certain high-risk subgroups had limited statistical power due to small numbers of patients and events, potentially obscuring any statistically significant differences. Larger studies or an individual patient data meta-analysis may help address these limitations. Finally, this study did not have a central adjudication of radiographic imaging and relied on the individual reporting of each center, which might have led to an inaccurate assessment of dissection features such as degree of stenosis or presence of intraluminal thrombus in some patients.

## Conclusion

In this post-hoc analysis of the STOP-CAD study, early presentation from first dissection symptom onset, acute infarct on baseline imaging, occlusive dissection, and prior history of ischemic stroke were identified as factors associated with a lower risk of subsequent ischemic stroke among patients with CAD. Among these high-risk subgroups, patients with occlusive dissection demonstrated a reduced stroke risk from anticoagulation. These findings can inform the design of future randomized trials and meta-analyses in order to validate the effectiveness of anticoagulants versus antiplatelets in reducing the risk of subsequent strokes in CAD patients, particularly those with occlusive dissection.

## Acknowledgements

None. This study was not funded.

## Author Contributions

Daniel Mandel, Liqi Shu, and Shadi Yaghi wrote the first draft of the manuscript, and all the authors reviewed and contributed to subsequent versions.

## References

1. Bejot Y, Daubail B, Debette S, Durier J, Giroud M. Incidence and outcome of cerebrovascular events related to cervical artery dissection: the Dijon Stroke Registry. Int J Stroke. Oct 2014;9(7):879–82. doi:10.1111/ijs.12154

2. Putaala J, Metso AJ, Metso TM, et al. Analysis of 1008 consecutive patients aged 15 to 49 with first-ever ischemic stroke: the Helsinki young stroke registry. Stroke. Apr 2009;40(4):1195–203. doi:10.1161/STROKEAHA.108.529883

3. Ekker MS, Verhoeven JI, Schellekens MMI, et al. Risk Factors and Causes of Ischemic Stroke in 1322 Young Adults. Stroke. Feb 2023;54(2):439–447. doi:10.1161/STROKEAHA.122.040524

4. Keser Z, Chiang CC, Benson JC, Pezzini A, Lanzino G. Cervical Artery Dissections: Etiopathogenesis and Management. Vasc Health Risk Manag. 2022;18:685–700. doi:10.2147/VHRM.S362844

5. Markus HS, Hayter E, Levi C, Feldman A, Venables G, Norris J. Antiplatelet treatment compared with anticoagulation treatment for cervical artery dissection (CADISS): a randomised trial. Lancet Neurol. Apr 2015;14(4):361–7. doi:10.1016/S1474-4422(15)70018-9

6. Kennedy F, Lanfranconi S, Hicks C, et al. Antiplatelets vs anticoagulation for dissection: CADISS nonrandomized arm and meta-analysis. Neurology. Aug 14 2012;79(7):686–9. doi:10.1212/WNL.0b013e318264e36b

7. Engelter ST, Traenka C, Gensicke H, et al. Aspirin versus anticoagulation in cervical artery dissection (TREAT-CAD): an open-label, randomised, non-inferiority trial. Lancet Neurol. May 2021;20(5):341–350. doi:10.1016/S1474-4422(21)00044-2

8. Compter A, Schilling S, Vaineau CJ, et al. Determinants and outcome of multiple and early recurrent cervical artery dissections. Neurology. Aug 21 2018;91(8):e769–e780. doi:10.1212/WNL.0000000000006037

9. Touze E, Gauvrit JY, Moulin T, et al. Risk of stroke and recurrent dissection after a cervical artery dissection: a multicenter study. Neurology. Nov 25 2003;61(10):1347–51. doi:10.1212/01.wnl.0000094325.95097.86

10. Kloss M, Grond-Ginsbach C, Ringleb P, Hausser I, Hacke W, Brandt T. Recurrence of cervical artery dissection: An underestimated risk. Neurology. Apr 17 2018;90(16):e1372–e1378. doi:10.1212/WNL.0000000000005324

11. Gensicke H, Ahlhelm F, Jung S, et al. New ischaemic brain lesions in cervical artery dissection stratified to antiplatelets or anticoagulants. Eur J Neurol. May 2015;22(5):859-65, e61. doi:10.1111/ene.12682

12. Yaghi S, Engelter S, Del Brutto VJ, et al. Treatment and Outcomes of Cervical Artery Dissection in Adults: A Scientific Statement From the American Heart Association. Stroke. Mar 2024;55(3):e91–e106. doi:10.1161/STR.0000000000000457

13. Menon R, Kerry S, Norris JW, Markus HS. Treatment of cervical artery dissection: a systematic review and meta-analysis. J Neurol Neurosurg Psychiatry. Oct 2008;79(10):1122–7. doi:10.1136/jnnp.2007.138800

14. Yaghi S, Shu L, Mandel DM, et al. Antithrombotic Treatment for Stroke Prevention in Cervical Artery Dissection: The STOP-CAD Study. Stroke. Feb 9 2024; doi:10.1161/STROKEAHA.123.045731

15. Kaufmann JE, Gensicke H, Schaedelin S, et al. Toward Individual Treatment in Cervical Artery Dissection: Subgroup Analysis of the TREAT-CAD Randomized Trial. Ann Neurol. Feb 16 2024; doi:10.1002/ana.26886

16. Dziewas R, Konrad C, Drager B, et al. Cervical artery dissection--clinical features, risk factors, therapy and outcome in 126 patients. J Neurol. Oct 2003;250(10):1179–84. doi:10.1007/s00415-003-0174-5

17. Klijn CJ, Kappelle LJ, Algra A, van Gijn J. Outcome in patients with symptomatic occlusion of the internal carotid artery or intracranial arterial lesions: a meta-analysis of the role of baseline characteristics and type of antithrombotic treatment. Cerebrovasc Dis. 2001;12(3):228–34. doi:10.1159/000047708

18. Liberman AL, Zandieh A, Loomis C, et al. Symptomatic Carotid Occlusion Is Frequently Associated With Microembolization. Stroke. Feb 2017;48(2):394–399. doi:10.1161/STROKEAHA.116.015375

